# Speech-Based Markers in Paediatric ADHD: A Longitudinal Case-Control Study of Voice Features and Medication Effects

**DOI:** 10.64898/2026.03.25.26348708

**Authors:** Rachel Bamberger, Gianna Kuhles, Leon D. Lotter, Juergen Dukart, Kerstin Konrad, Thomas Günther, Michael Siniatchkin, Michael Fuchs, Georg von Polier

**Author notes:** Corresponding author: Correspondence to Georg von Polier and Rachel Bamberger.

## Abstract

**Background:** Diagnosis and treatment monitoring of attention-deficit/hyperactivity disorder (ADHD) largely rely on subjective assessments, highlighting the need for objective markers. Voice features and speech embeddings represent promising candidates for such markers, as they may capture alterations in speech production relevant to ADHD. However, it remains unclear which speech features are most informative for distinguishing ADHD and monitoring treatment effects, and which speech tasks most reliably elicit such differences.

**Methods:** Twenty-seven children with ADHD and 27 age-matched neurotypical controls completed six speech tasks across two study visits. Children with ADHD were unmedicated at baseline (first visit) and were assessed under prescribed methylphenidate treatment at follow-up, whereas controls underwent repeated assessment without intervention. Established acoustic voice features (eGeMAPS) and high-dimensional speech embeddings (WavLm, Whisper) were extracted and analysed using linear mixed models to examine baseline group differences and group-by-time interaction effects reflecting medication-associated change patterns.

**Results:** At baseline, children with ADHD differed significantly from controls in frequency, spectral, and temporal voice features, characterized by lower and more variable pitch, altered spectral properties, and reduced rhythmic stability. Group-by-time interaction effects indicated medication-associated modulation in the ADHD group, including reduced loudness variability and increased precision of vowel articulation at follow-up, changes not observed in controls. Speech embeddings revealed additional baseline and interaction effects beyond established acoustic features. Free speech tasks, particularly picture description, yielded the most robust and consistent effects.

**Conclusion:** Children with ADHD differed from neurotypical controls in vocal features at baseline and showed distinct longitudinal change patterns consistent with medication-related change. These findings support further investigation of speech-based measures as candidate digital phenotypes and potential digital biomarkers in ADHD, with picture description emerging as a particularly promising task for future clinical assessment protocols.

## 1 Introduction

Diagnosis and treatment evaluation of attention-deficit/hyperactivity disorder (ADHD) rely largely on subjective assessments by clinicians, parents, and patients [1]. Limitations thereof include high observer bias as well as insufficient validity or reliability [2]. This has led to a broad consensus on the need for more objective markers [1, 3, 4]. Objective markers may provide additional information for diagnosis and facilitate more personalised ADHD treatment [2, 5].

Speech production is one of the most complex and elaborate motor outputs of the brain [6]. The voice conveys information beyond plain content and could therefore provide non-invasive insights into neurodevelopmental disorders [7]. Research into voice features as biomarkers within psychiatry has steadily increased, and their use appears promising [8, 9].

Deficits in neuropsychological mechanisms, including motor control or cognitive processes, could alter speech patterns. For example, the necessary synchronisation between motor output and sensory perception required for speech production seems to be different in individuals with ADHD, leading to a faster speaking rate and alternating motion ranges [10]. Furthermore, deficits in inhibitory control in ADHD seem to impact the speech production system requiring this control mechanism to prevent or correct errors while speaking [11]. Research on voice in children with ADHD shows more hyperfunctional vocal behaviour, i.e. speaking with a louder voice or applying more pressure [12] as well as more hoarseness, straining and breathiness [13, 14]. Our previous research in clinical and population samples has demonstrated an association between voice features, such as pitch and intensity, and ADHD symptoms in children and adults [15, 16]. Preliminary evidence suggests that voice features may be sensitive to pharmacological modulation with methylphenidate (MPH) supporting their potential utility for treatment monitoring. Specifically, changes in fundamental frequency (F0) [17] and jitter (an acoustic parameter of F0 stability) [18] were observed following MPH administration.

Established voice features allow hypothesis-driven investigation of specific acoustic parameters, but do not capture other speech features known to be compromised in ADHD. Speech embeddings provide an additional, exploratory approach by encoding high-dimensional speech representations that integrate multiple aspects of speech, including (para)linguistic cues, prosody, and temporal dynamics [19, 20]. In this sense, embeddings can be used to explore whether speech contains additional relevant information beyond established interpretable voice features.

Importantly, both voice features and speech-embeddings depend critically on the speech context in which they are elicited. In clinical practice, multiple speech tasks are used to capture distinct vocal and speech information. Free speech paradigms, such as picture description, are the most commonly applied and highly standardised [21]. Other frequently used tasks include sustained phonation, diadochokinesia, and reading.

The present study aimed to evaluate the potential utility of speech-based measures in a clinical sample by (a) comparing voice features and speech embeddings between unmedicated children with ADHD and age-matched neurotypical controls and (b) examining longitudinal changes in these measures following MPH treatment in children with ADHD, compared with repeated assessments without treatment in controls. It is important to note that this study primarily focused on established voice features. As speech embeddings contain information that is not directly interpretable, analysis remained largely exploratory. Additionally, we systematically investigated which speech tasks were most sensitive to differences and changes in speech features, to inform future task selection. Based on the existing literature, we hypothesised that speech features contain information relevant to distinguishing between children with ADHD and neurotypical controls, as well as to detecting MPH treatment response. We also hypothesised that certain speech tasks are better at revealing potential differences or changes in speech than others. Due to the heterogeneity of previous findings, no directional hypotheses were formulated.

## 2 Methods

### 2.1 Study design and participants

We conducted a case-control study with a pre-post design. The data were acquired at the Department of *Child and Adolescent Psychiatry, Psychotherapy and Psychosomatics at RWTH Aachen University Hospital*. The study is in accordance with the Declaration of Helsinki, was approved by the *Ethics Committee of the Medical Faculty of RWTH Aachen University* (approval no. 032/20), and registered with the *German Clinical Trials Registry* (DRKS; registration no. DRKS00030766). Written informed consent was received from all parents and children, regardless of age.

We included children aged between 8 and 16 years who were either neurotypical or had a clinical ADHD diagnosis according to DSM-5 criteria and had been prescribed pharmacotherapy with MPH. Children with an IQ below 80 or a comorbid autism spectrum disorder were excluded. For the present study, boys who already underwent pubertal voice change (fundamental frequency below *C3*, i.e., 130.82 Hz [22]) were excluded from the analysis. Children with ADHD were recruited from within the clinic and local outpatient practices, neurotypical controls (NC) through public advertisements (flyers/mailing lists).

Participation involved two visits (baseline/follow-up) at our clinic, ideally scheduled about 8 weeks apart. These included an IQ estimation (if a full assessment was not available), neuropsychological and speech tasks, questionnaires and, for eligible and willing participants, an optional MRI session. For the present study, data from the speech tasks (≈ 20 minutes) and questionnaires were used, as described further below.

A total of 57 children participated in the study. Three males of the control group had already undergone voice change, so their data were excluded. The remaining 54 children (female = 31; male = 21, diverse = 2; ADHD = 27; NC = 27) had a mean age of 12.40 years (range: 8.08 - 16.92). Out of the whole sample, only two children with ADHD discontinued pharmacotherapy and were therefore not assessed at follow-up.

### 2.2 Measurements

#### 2.2.1 Demographic and health data

Demographic (age, gender) and health data were collected through interviews and questionnaires, diagnostic information (ADHD subtype and comorbidities according to ICD-10 criteria) was obtained from clinic records. To assess whether DSM-5 criteria for ADHD were met, parents completed the *Attention Deficit Hyperactivity Disorder* supplement of the *Schedule for Affective Disorders and Schizophrenia for School-Age Children—Present and Lifetime Version* (K-SADS-PL) [23]. Absence of a diagnoses in controls was confirmed by the parents and validated by evaluation of the K-SADS-PL.

At both visits, parents completed the long form of the German adaption of the *Conners 3^rd^ Edition-Parent* (Conners 3-P) [24]. This questionnaire is designed to assess symptoms of ADHD and their severity in children aged 6–18. For the present study, we used the values of children with ADHD on the *Inattention* (IA) and *Hyperactivity/Impulsivity* (HI) scales.

#### 2.2.2 Voice measurements

Voice measurements were conducted according to a standard operating procedure (SOP), which is guideline recommended [25, 26]. The SOP included detailed written instructions regarding equipment and setup, recording parameters, and verbal instructions for each task to ensure reproducible voice recordings (**Appendix S1**).

During data collection, the recording setup was updated in line with current guideline recommendations (**Appendix S2**). Consequently, ten participants (9 with ADHD) were recorded using the earlier setup. The initial setup employed a cardioid microphone (*Elgato Wave:1*) with a microphone-mouth distance between 30-40 cm. The final recording setup used an omnidirectional headset microphone (*Sennheiser HSP 2 EW 3*) positioned 2 cm from the corner of the mouth. All recordings were obtained at a sample rate of 48.000 Hz at 24-bit resolution and were stored in the *.wav* audio format [25].

Recordings were conducted in a quiet room with the child and the researcher seated opposite each other. During both visits, children were asked to complete a total of six speech tasks in German (**Table 1)**. The speech task protocol was slightly refined in the early stages of the data collection to reflect updated methodological recommendations. A five-second silent recording was added, to analyse ambient noise levels during each visit [26]. Two sustained phonation tasks (normal/loud voice) were included to enable reliable analysis of jitter values [27]. Additional recordings were not taken for the first 5 children (ADHD) who participated.

**Table 1.**
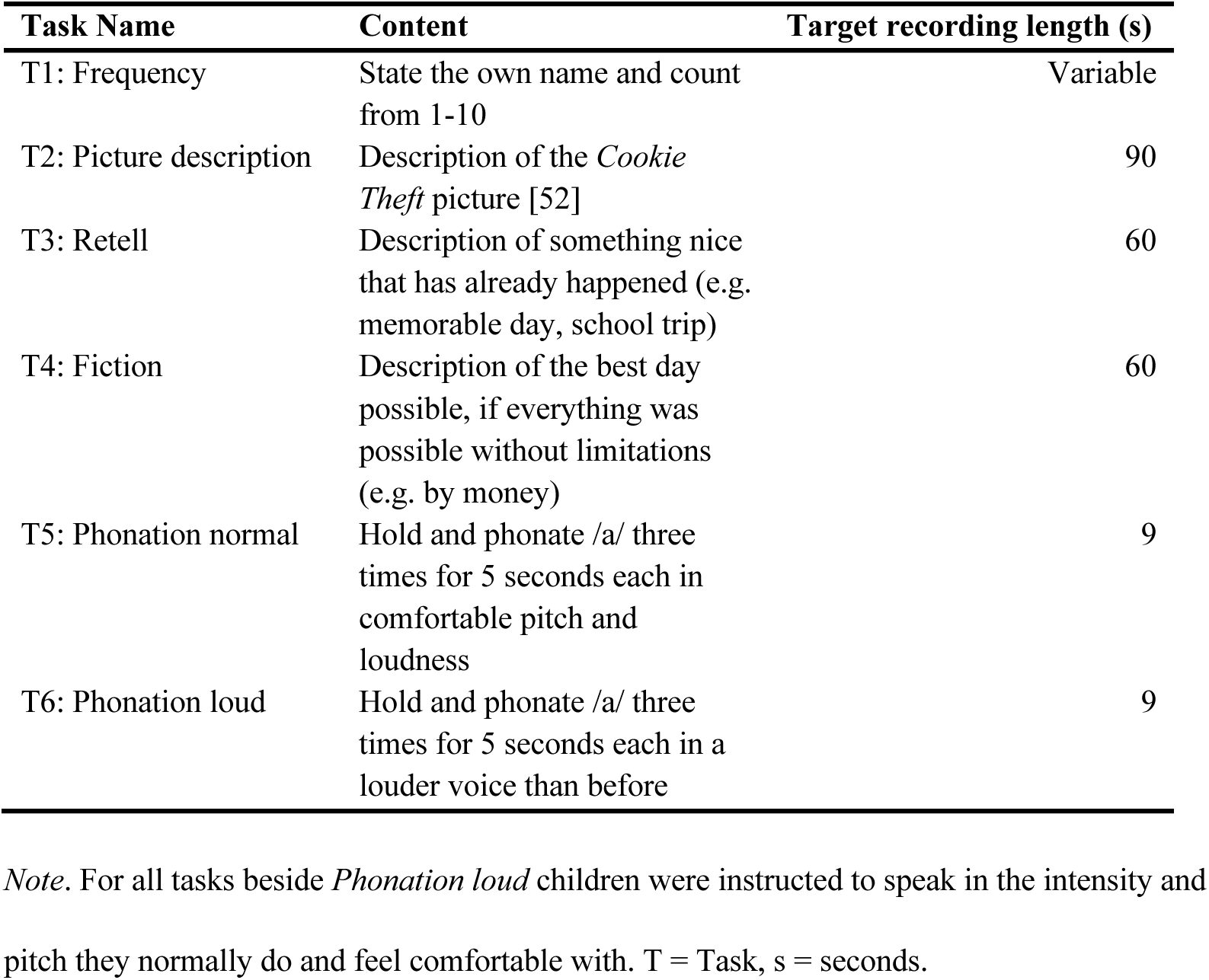
Name, content and target recording length of the six speech tasks for both visits.

| Task Name | Content | Target recording length (s) |
| --- | --- | --- |
| T1: Frequency | State the own name and count from 1-10 | Variable |
| T2: Picture description | Description of the *Cookie Theft* picture [52] | 90 |
| T3: Retell | Description of something nice that has already happened (e.g. memorable day, school trip) | 60 |
| T4: Fiction | Description of the best day possible, if everything was possible without limitations (e.g. by money) | 60 |
| T5: Phonation normal | Hold and phonate /a/ three times for 5 seconds each in comfortable pitch and loudness | 9 |
| T6: Phonation loud | Hold and phonate /a/ three times for 5 seconds each in a louder voice than before | 9 |

*Note.* For all tasks beside *Phonation loud* children were instructed to speak in the intensity and pitch they normally do and feel comfortable with. T = Task, s = seconds.

#### 2.2.3 Voice features

Voice features were automatically extracted from the speech recordings using the standardised and widely used *eGeMAPS* feature set [28], which is implemented in the *openSMILE* toolkit [29]. The eGeMAPS set comprises 88 theoretically grounded voice features, extracted separately from each task. These features are derived from four voice-domains: Frequency (e.g. pitch), Energy/amplitude (e.g. loudness), Spectral (e.g. Mel-Frequency Cepstral Coefficients), and Temporal (e.g. mean length of voiced regions).

#### 2.2.4 Speech embeddings

Speech embeddings were automatically extracted using the *WavLM* [30] and *Whisper* [31] base encoders, resulting in 768-dimensional representations per child, task, and visit. These transformer-based models generate high-dimensional representations of speech that capture e.g. (para)linguistic, prosodic, and temporal characteristics. Both models are trained on large, varied datasets, so their embeddings reflect multidimensional speech dynamics and may reveal subtle speech changes relevant for clinical or behavioural characterisation [19].

### 2.3 Statistical Analysis

#### 2.3.1 Data preparation and quality control

The audio recordings were preprocessed using *Python* (3.9) and custom scripts that leveraged libraries including *librosa* (0.11) [32], *praat-parselmouth* (0.4) [33], and *webrtcvad* (2.0). All files were resampled to 48.000 kHz in a 16-bit format to comply with the preprocessing packages and feature extraction methods. The recordings were cut according to the respective task **(Appendix S3)** and the targeted recording length **(Table 1)**. Each recording’s audio wave was visually inspected and a minimum of 10% of recordings from each task was fully listened to and checked for inconsistencies. Quality control of ambient noise levels and signal-to-noise-ratio (SNR) was performed using the recordings of silence (ambient noise) and phonation normal (SNR). All recordings met the guideline described quality criteria [26, 34] **(Appendix S4)**. Following preprocessing and quality control, audio files were passed to the feature extraction pipelines using Python scripts as described above.

#### 2.3.2 Linear mixed models

We used linear mixed models (LMMs) to assess effects of group at baseline as well as group-by-time interactions on voice features. All variables were z-standardized independently across subjects and sessions. Intensity-based features (i.e., loudness-related) were set to missing (*NaN)* for recordings without the headset microphone as these features are sensitive to recording distance and equipment. Separate LMMs were fitted for each of the 88 voice features extracted from each task. Features exhibiting zero variance were excluded from analysis, resulting in a total of 512 models. Models were specified in a 2×2 design with group (controls/ADHD; reference: controls) and time (baseline/follow-up; reference: baseline) as main effects, their interaction, as well as age at baseline and gender (m/w/d) as covariates [35]. Using this design, the main effect of group can be interpreted as the difference between ADHD and controls at baseline. We interpreted significant interaction effect as indication for a difference in changes observed over time. Random intercepts accounted for within-subject variability. Features were excluded if they were structurally invalid for the task (i.e. unvoiced features in phonation tasks) or showed zero variance. LMMs were fitted using the python *statsmodels* (0.14.5) package and model fit was optimised using a standard procedure (*Powell* method, 1000 iterations). Multiple testing was corrected task-wise using the False Discovery Rate (FDR; Benjamini-Hochberg procedure). Results with FDR-corrected p-values (q-values) < 0.05 were considered statistically significant. To assess robustness to outliers that may occur due to the high inter-and intrapersonal variability of voice [36], the LMM analyses were repeated using ranked data, resembling a rank-based Wilcoxon-like procedure [37]. As LMM estimates do not typically allow for standardised effect size measures, the fixed-effects regression coefficient (β) was used as a measure of effect magnitude. Previously done z-transformation ensured that the coefficients were comparable among predictors. Additionally, we reran the analyses excluding three children with ADHD assessed during their medication pause and robust interaction effects were re-tested by including the *time between sessions* as an additional covariate. Post-hoc analyses using paired t-tests on session-to-session change scores were conducted within the ADHD group to explore significant group-by-time interaction effects. Clinical relevance was examined by correlating significant baseline voice features with parent-rated ADHD raw symptom scores. Additionally, linear regression models were used to test whether changes in ADHD symptoms (IA and HI) predicted changes in voice features, adjusting for age and gender. Given the small sample size and the exploratory nature of the study, no correction for multiple comparisons was applied.

Speech embeddings were analysed using LMMs largely analogous to those applied to voice features. Due to the high dimensionality of the embeddings (768 dimensions per model), task-specific *Principal Component Analysis* (PCA) was conducted on baseline data. For each task, the first ten principal components (PCs) were retained and used for subsequent analyses. Using a fixed number of components ensured comparable dimensions while maintaining consistency for the subsequent analysis. For each of the two encoder models, 10 PCs were entered as fixed effects in the LMM alongside baseline age and gender. This resulted in 60 separate LMMs for each encoder (10 PCs by six tasks).

To assess whether observed effects of speech embeddings were independent of established voice features, PCs showing significant baseline or interaction effects were reanalysed including specific voice features as covariates (mean F0, standard deviation of MFCC1, and standard deviation of voiced-segment length).

## 3 Results

### 3.1 Demographic and clinical characteristics

**Table 2** shows demographic and clinical characteristics of the 54 included participants. The groups did not differ significantly in age (*t* = 1.14, *p* =.260) or gender (χ² = 2.08, *p* =.353) distributions at baseline. ADHD symptoms (Conners 3-P) significantly decreased from baseline to follow-up in the ADHD group (Inattention (IA): *t*(23) = 5.63, *p* <.001; Hyperactivity/Impulsivity (HI): *t*(23) = 3.98, *p* =.001), whereas no significant change was observed in controls (IA: *t*(23) =-1.85, *p* = 0.08; HI: *t*(23) = 0.13, *p* = 0.90). 47 participants had voice data available for both visits (see **Appendix S5** for information on missing sessions). The mean interval between visits was 11.93 weeks (SD = 6.18) for children with ADHD and 8.42 weeks (SD = 0.82) for controls. The longer and more variable interval in the ADHD group reflects differences in the timing of pharmacotherapy initiation among drug-naïve participants. Details on recording equipment, the number of completed tasks per visit, and the average duration of preprocessed recordings per task are provided in **Table S1**.

**Table 2.** Demographic and clinical information for both visits.

| Characteristic | Group | n (%) (Baseline) | Mean/SD (Baseline) | n (Follow-Up) | Mean/SD (Follow-Up) |
| --- | --- | --- | --- | --- | --- |
| **Age** | ADHD | 27 | 12.79 (2.65) | 25 | 13.04 (2.77) |
|  | Control | 27 | 12.01 (2.38) | 27 | 12.17 (2.38) |
| **Gender** |  |  |  |  |  |
| Male | ADHD | 10 | - | - | - |
|  | Control | 11 | - | - | - |
| Female | ADHD | 15 | - | - | - |
|  | Control | 16 | - | - | - |
| Divers | ADHD | 2 | - | - | - |
|  | Control | 0 | - | - | - |
| **ADHD Subtype** | ADHD |  |  |  |  |
| Combined |  | 18 | - | - | - |
| Inattentive |  | 8 | - | - | - |
| Hyperactive-Impulsive |  | 1 | - | - | - |
| **Conners3-P** |  |  |  |  |  |
| Inattention | ADHD | 25 | 17.72 (5.52) | 24 | 11.79 (6.65) |
|  | Control | 26 | 4.12 (3.28) | 25 | 5.24 (4.13) |
| Hyperactivity/ Impulsivity | ADHD | 25 | 17.20 (10.05) | 24 | 11.04 (8.59) |
|  | Control | 26 | 4.08 (4.13) | 25 | 3.88 (4.48) |
| **Comorbid diagnosis** | ADHD | 22 (81.48) | - | - | - |
| Depressive Disorders |  | 13 (48.15) | - | - | - |
| Anxiety Disorders |  | 12 (44.44) | - | - | - |
| Trauma-related Disorders |  | 4 (14.81) | - | - | - |
| Gender Dysphoria |  | 2 (7.41) | - | - | - |
| Eating Disorders |  | 2 (7.41) | - | - | - |
| Specific Learning Disorder |  | 2 (7.41) | - | - | - |
| Somatic Symptom Disorder |  | 2 (7.41) | - | - | - |
| Conduct Disorders |  | 1 (3.70) | - | - | - |
| Communication Disorders |  | 1 (3.70) | - | - | - |
| Elimination Disorders |  | 1 (3.70) | - | - | - |

*Note.* SD = Standard deviation; Conners3-P = Conners 3rd Edition-Parent (long form). Comorbid diagnoses were limited to psychiatric and behavioural disorders classified in the DSM-5. Comorbid diagnoses were not mutually exclusive, meaning that children could meet the criteria for multiple additional diagnoses.

#### Medication

Children with ADHD were unmedicated at the baseline visit. Twenty-two patients were initially drug-naïve and prescribed MPH according to routine clinical care. Titration, timing of initiation, and dosage were individualised in line with current guidelines. Three participants had paused prior MPH treatment before baseline (treatment duration before study entry: 5 years, 10 months, 4 months; pause duration before follow-up: 41, 44 and 51 days).

Following the baseline assessment, MPH treatment was initiated/resumed according to the individual treatment plan. Participants were expected to receive continuous treatment for approximately eight weeks until follow-up. At follow-up, children with ADHD were assessed while taking their individually prescribed MPH medication. Twenty-three children received an extended-release formulation, one an immediate-release formulation, and one a combination of both (**Table S2** for details). Voice recordings were conducted on medication, on average 3 h 51 min (SD = 1 h 13 min) after the last intake.

### 3.2 Voice features

#### 3.2.1 Baseline differences

Six unique voice features across three tasks showed significant baseline group differences after FDR correction **(Table 3**, **Figure 1)**. These features were predominantly pitch-related and included measures of F0, capturing both mean level and variability. The values of these features indicated that children with ADHD had lower voices that varied more during speech and therefore seemed less stable. Spectral features also contained discriminative information, particularly the first and second Mel-frequency cepstral coefficients (MFCCs), which characterize spectral shape such as voice timbre. These parameters showed that voices of children with ADHD were brighter/tenser (MFCC1), and less variable in timbre (MFCC2), meaning the tone colour fluctuated less. Additionally, the standard deviation of voiced-segment length, an indicator of rhythmic stability, differentiated between the groups, suggesting that the voices of children with ADHD were less rhythmically stable. Residuals of the LMM were approximately normally distributed. Robustness checks using ranked-transformed data to mitigate the influence of outliers yielded mostly consistent results with respect to effect estimates and signs. Comparable results were obtained when excluding the three children with ADHD who were assessed during a medication pause. Additionally, various features displayed significant group differences not surviving correction for multiple comparisons **(Table S3)**.

**Figure 1.**
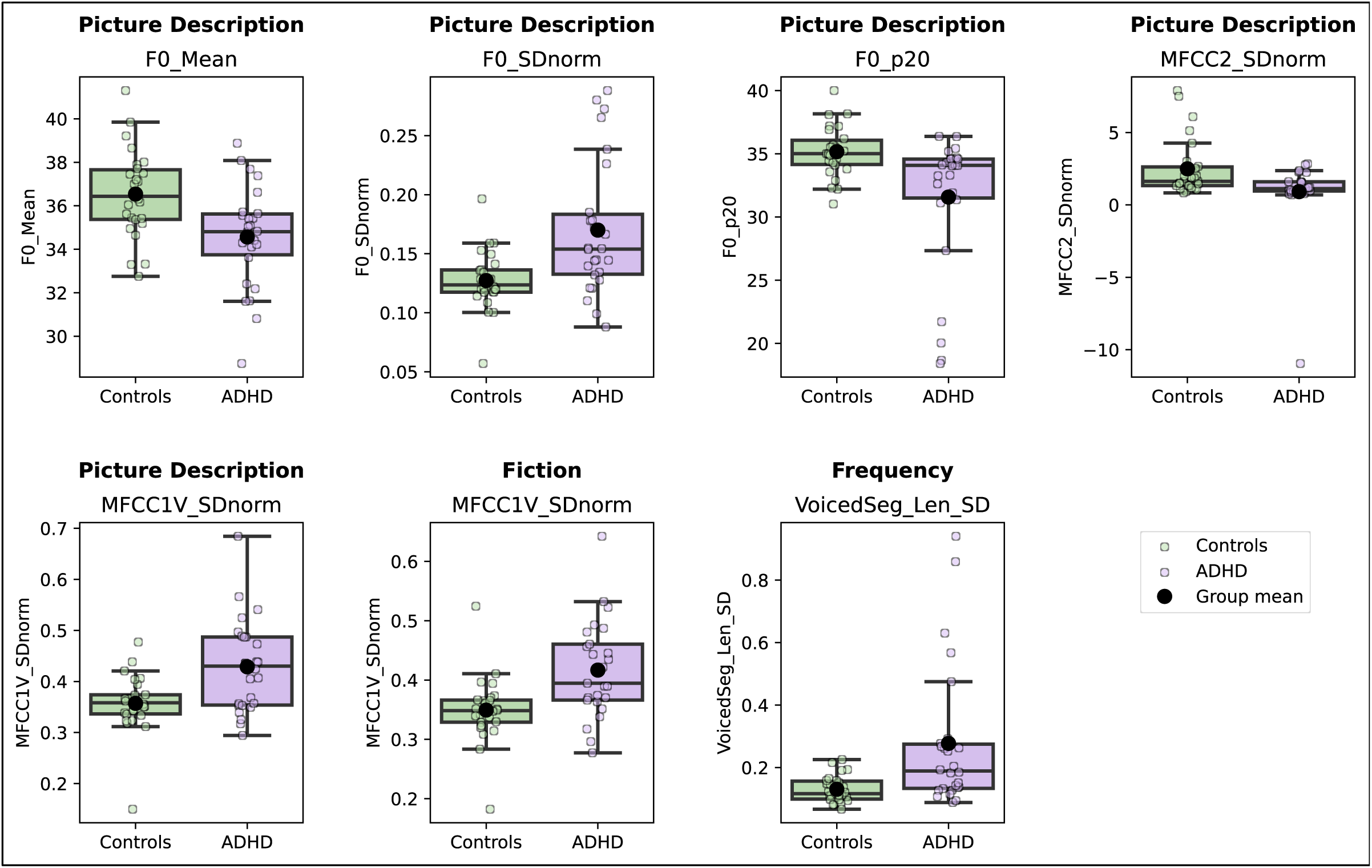
FDR-corrected significant voice features at baseline (linear mixed model) *Note.* All values represent the raw values of the significant effects found in the linear mixed model analysis. F0_Mean = mean pitch (fundamental frequency); F0_SDnorm = normalized pitch variability; F0_p20 = 20th percentile pitch; MFCC2_SDnorm = normalized variability of 2nd Mel-frequency cepstral coefficient; MFCC1V_SDnorm = normalized variability of 1st MFCC in voiced regions; VoicedSeg_Len_SD = standard deviation of voiced segment lengths.

**Table 3.**
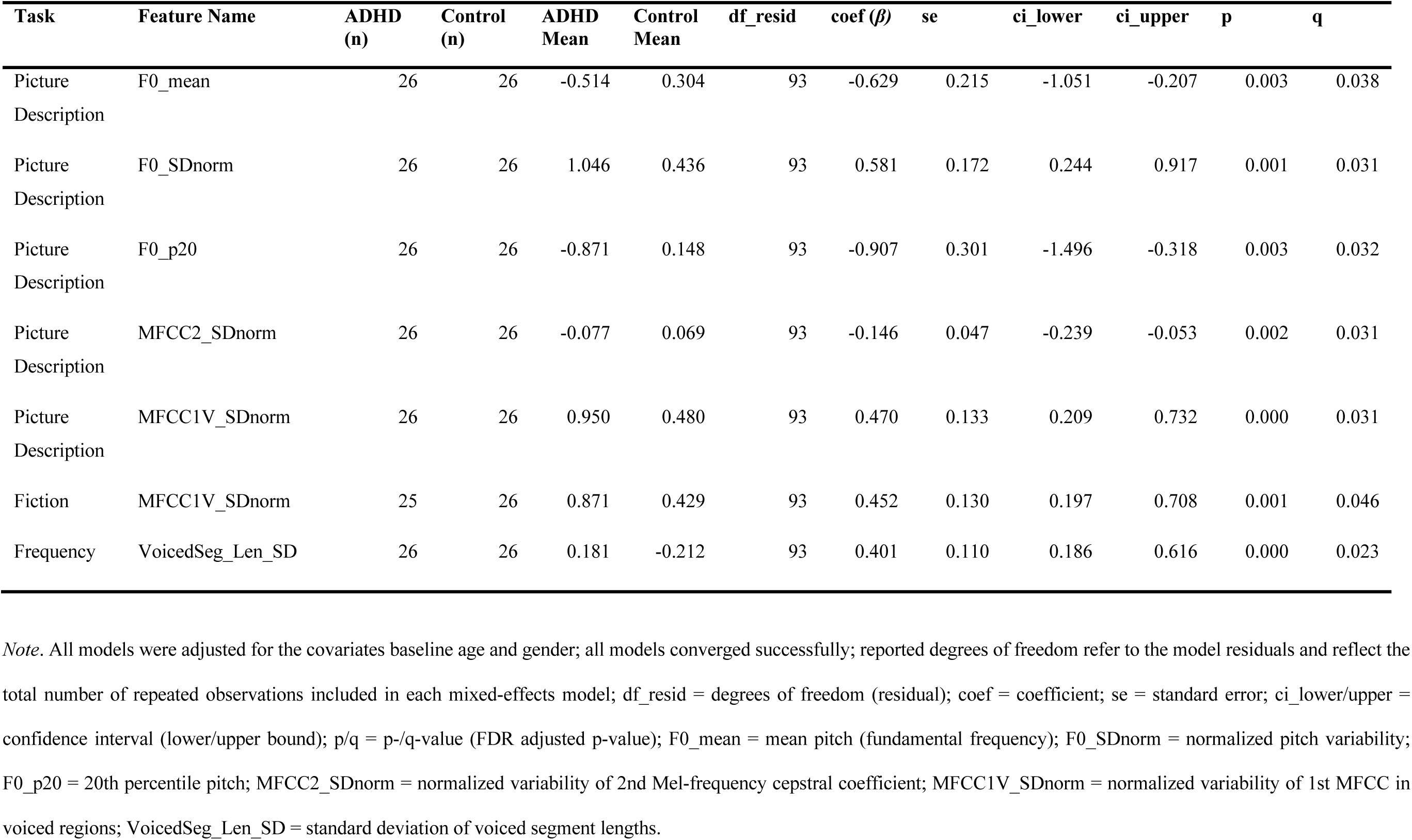
FDR-corrected significant voice features at baseline (linear mixed model)

| Task | Feature Name | ADHD<br>(n) | Control<br>(n) | ADHD<br>Mean | Control<br>Mean | df_resid | coef ($\beta$) | se | ci_lower | ci_upper | p | q |
| --- | --- | --- | --- | --- | --- | --- | --- | --- | --- | --- | --- | --- |
| Picture<br>Description | F0_mean | 26 | 26 | -0.514 | 0.304 | 93 | -0.629 | 0.215 | -1.051 | -0.207 | 0.003 | 0.038 |
| Picture<br>Description | F0_SDnorm | 26 | 26 | 1.046 | 0.436 | 93 | 0.581 | 0.172 | 0.244 | 0.917 | 0.001 | 0.031 |
| Picture<br>Description | F0_p20 | 26 | 26 | -0.871 | 0.148 | 93 | -0.907 | 0.301 | -1.496 | -0.318 | 0.003 | 0.032 |
| Picture<br>Description | MFCC2_SDnorm | 26 | 26 | -0.077 | 0.069 | 93 | -0.146 | 0.047 | -0.239 | -0.053 | 0.002 | 0.031 |
| Picture<br>Description | MFCC1V_SDnorm | 26 | 26 | 0.950 | 0.480 | 93 | 0.470 | 0.133 | 0.209 | 0.732 | 0.000 | 0.031 |
| Fiction | MFCC1V_SDnorm | 25 | 26 | 0.871 | 0.429 | 93 | 0.452 | 0.130 | 0.197 | 0.708 | 0.001 | 0.046 |
| Frequency | VoicedSeg_Len_SD | 26 | 26 | 0.181 | -0.212 | 93 | 0.401 | 0.110 | 0.186 | 0.616 | 0.000 | 0.023 |

*Note.* All models were adjusted for the covariates baseline age and gender; all models converged successfully; reported degrees of freedom refer to the model residuals and reflect the total number of repeated observations included in each mixed-effects model; df\_resid = degrees of freedom (residual); coef = coefficient; se = standard error; ci\_lower/upper = confidence interval (lower/upper bound); p/q = p-/q-value (FDR adjusted p-value); F0\_mean = mean pitch (fundamental frequency); F0\_SDnorm = normalized pitch variability; F0\_p20 = 20th percentile pitch; MFCC2\_SDnorm = normalized variability of 2nd Mel-frequency cepstral coefficient; MFCC1V\_SDnorm = normalized variability of 1st MFCC in voiced regions; VoicedSeg\_Len\_SD = standard deviation of voiced segment lengths.

The picture description task, followed by fiction and retell tasks, appeared to best elicit baseline differences. Features derived from the picture description task yielded the largest absolute *β*-values, indicating generally stronger effects, resulting in several significant voice features under both uncorrected (n = 5) and FDR-corrected (n = 5) conditions.

#### 3.2.2 Interaction effects

Two voice features from separate tasks exhibited significant group-by-time effects after FDR correction, indicating that these parameters changed differently between visits in children with ADHD compared to controls **(Table 4**, **Figure 2)**. Notably, the picture description task revealed the strongest interaction effect: The mean bandwidth of the first formant (F1) increased in the ADHD group, largely converging with the comparably stable control values. This indicates a more pronounced change in vowel articulation for children with ADHD and is consistent with clearer articulation over time. The derived results of the fiction task showed that the ADHD group exhibited decreased loudness variability, suggesting a more stable energy pattern over time, while the control group showed greater variability. Residuals were approximately normally distributed. The interaction effects remained stable in ranked-data LMMs and while excluding the three children with paused medication. Additionally post-hoc sensitivity analyses with adjustment for the time between visits, that differed between the two groups, produced similar results, with the group × time interaction effects remaining unchanged in terms of direction and significance. Features not surviving correction for multiple comparisons are reported in **Table S4**.

**Figure 2.**
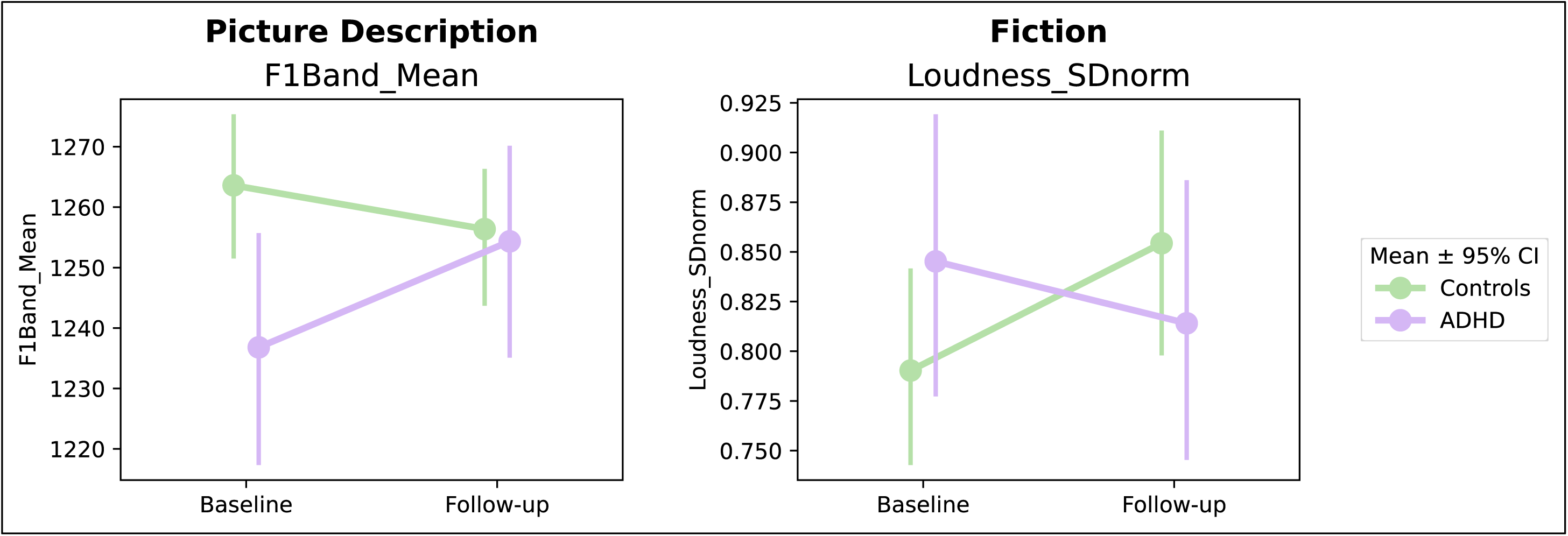
Raw values of FDR-corrected significant Group × Time interaction effects in voice features (linear mixed model) *Note.* All values represent the raw values of the significant effects found in the linear mixed model analysis. Children with ADHD were under medication at follow-up. F1Band_mean = mean bandwidth of the first formant; Loudness_SDnorm = normalized variability of loudness; CI = confidence interval.

**Table 4.** FDR-corrected significant Group × Time interaction effects in voice features (linear mixed model)

| Task | Feature Name | ADHD<br>Baseline/<br>Follow-Up | Control<br>Baseline/<br>Follow-Up | ADHD Mean<br>Baseline/<br>Follow-Up | Control Mean<br>Baseline/<br>Follow-Up | df_resid | coef ($\beta$) | se | ci_lower | ci_upper | p | q |
| --- | --- | --- | --- | --- | --- | --- | --- | --- | --- | --- | --- | --- |
| Picture<br>Description | F1_bandwidth_mean | 26/24 | 26/24 | -0.393/-0.219 | -0.134/-0.201 | 93 | 0.276 | 0.078 | 0.123 | 0.429 | 0.000 | 0.037 |
| Fiction | Loudness_SDnorm | 17/15 | 24/23 | 0.684/0.582 | 0.527/0.716 | 72 | -0.353 | 0.097 | -0.543 | -0.163 | 0.000 | 0.024 |

*Note.* All models were adjusted for the covariates baseline age and gender; all models converged successfully; df\_resid = degrees of freedom (residual); coef = coefficient; se = standard error; ci\_lower/upper = confidence interval (lower/upper bound); p/q = p-/q-value (FDR adjusted p-value); F1\_bandwidth\_mean = mean bandwidth of the first formant; Loudness\_SDnorm = normalized variability of loudness.

Post-hoc analysis on the change scores within the ADHD group confirmed significant changes over time with moderate within-group effect sizes (Bandwidth F1: *t*(23) = 3.54, *p* =.002, dz = 0.72; Loudness: *t*(14) =-2.30, *p* =.038, dz =-0.59).

#### 3.2.3 Clinical Relevance

##### Correlation voice features and ADHD symptom ratings

Associations between baseline voice features and parent-rated ADHD symptoms were generally small and not statistically significant. However, mean f0 (Fiction), showed a moderate positive correlation with hyperactivity/impulsivity (HI) values (*ρ* =.407, *p* =.048) consistent with the direction of the baseline group differences identified in the LMM analyses.

##### Linear Regression Model using change scores

Linear regression analyses were conducted using voice features that showed a robust interaction effect (**Table 4**). Within the ADHD group, changes in inattentive (IA) symptoms were significantly associated with changes in the mean bandwidth of the first formant (F1) (β = −0.032, SE = 0.014, p =.034). This indicates that a greater reduction in IA symptoms corresponds to larger increase in F1 bandwidth and thus more pronounced vowel articulation. Changes in HI symptoms were not associated with F1 bandwidth (p =.272). For loudness variability, neither symptom dimension showed significant associations with voice changes (IA: *p* =.523; HI: *p* =.230).

### 3.3 Speech Embeddings

Separate LMMs for each PC revealed significant baseline and group-by-time interaction effects for both WavLM and Whisper embeddings, after adjustment for baseline age, gender and selected voice features [f0 mean, MFCC1 standard deviation (SD), voiced-segment length (SD)]. For the WavLM model, two interaction effects remained significant after FDR correction; the other effects did not survive multiple comparison analysis **(Table S5)**. Whisper embeddings showed four baseline effects and one interaction effect that survived FDR correction **(Table S6)**. Taken together, these findings suggest that speech embeddings capture variance not accounted for by theoretically defined voice features and may provide complementary information on ADHD–control differences at baseline and medication-related changes over time. Across tasks, the first 10 principal components explained on average 76.82% (WavLm) / 65.57% (Whisper) of the variance. Explained variance for each PC and task are reported in **Table S7** (WavLM) and **Table S8** (Whisper).

## 4 Discussion

This study represents a proof-of-concept investigation of voice-based markers in a clinically characterised ADHD sample. The primary aim was to evaluate the suitability of voice features to distinguish between children with ADHD and controls and to capture treatment-related changes. Our main findings demonstrate that voice features robustly capture both group differences between children with ADHD and neurotypical controls and potential medication-related changes in children with ADHD, in a task-dependent manner. Exploratory analyses using speech embeddings suggest that the speech signal may contain additional information beyond established voice features, motivating more targeted investigations of, e.g., (para)linguistic features in future studies.

Several voice features differed significantly in children with ADHD compared to controls. Unmedicated children with ADHD had a lower, more variable voice pitch. At the same time, the colour of their voices seemed to be brighter and more tense, which could be perceived as pressed. This is in line with previous findings, describing a lower but more strained voice in children with ADHD [13, 14]. A strained voice may result from excessive vocal effort, which can be caused by altered motor regulation accompanied by heightened muscle tone in ADHD [38]. Furthermore, children with ADHD appear to exhibit more phonotraumatic behaviour, which involves placing greater mechanical stress on the vocal folds [39]. From a neuropsychological perspective, hypoarousal [40] may weaken top-down regulatory control necessary for vocal motor output and pitch regulation, contributing to increased pitch variability. Children with ADHD spoke less rhythmically during the counting task, which involved counting from one to ten. Counting is repetitive and therefore has a rhythm of its own. Children with ADHD may struggle to generate an internal beat [41] and have difficulties with the precise timing of motor tasks [42], which may explain this finding. We also found that the voice in both groups changed differently over time. After medication, children with ADHD showed a clearer articulation and a more stable loudness pattern. ADHD has been linked to changes in the brain’s dopamine signalling [43], which could affect voice production [44]. The medication’s effects include altering central dopamine activity [45], which could positively influence speech production. Controls displayed a reversed pattern for loudness variability, a speculative interpretation is increased comfort or familiarity at follow-up, though this cannot be determined from the current data. For articulation precision, the values almost merged from the first to the second visit. Given the naturalistic design, the effects of medication and time cannot be fully disentangled. However, the absence of comparable changes in the control group suggests that the changes observed are medication related.

We found multiple significant baseline and interaction effects for the speech embeddings. Given their limited interpretability, especially following necessary dimensionality reduction, we avoid a discussion of the meaning of significant embeddings. However, their training objectives could provide indirect clues about the types of speech information they encode (predominantly acoustic-prosodic vs. linguistic-phonetic). While our results demonstrate the potential of embedding-based analysis of speech data in ADHD, future research should examine which (para)linguistic aspects are captured by embeddings. This is crucial as underlying neuropsychological processes, such as emotional self-regulation and impulsivity [46] as well as inhibitory control [11], may contribute to altered (para)linguistic patterns in ADHD. Further exploration of the capability of the two models to capture baseline (Whisper) vs. longitudinal (WavLM) alterations would also be valuable. Combining acoustic features with speech embeddings in machine learning models may further improve performance [47].

The final stated goal of this study was to examine speech tasks in terms of their ability to discriminate between children with ADHD and controls, to provide prior knowledge for future task design. The picture description task yielded the most robust features, likely because children produce similar content, while differing subtly in prosodic delivery. This dual benefit makes description tasks suitable to elicit connected speech, which can provide clinical information as previous research has shown [48, 49]. The fiction and retell tasks also appear suitable. They require the children to either generate content freely and think outside of the box, or to retrieve, hold and manipulate prior knowledge to verbalise it. These tasks require cognitive resources enabled by e.g. executive functions (EF) like cognitive flexibility and working memory [50]. Speech and articulation also rely on coordinated neural networks [6] and higher order EF [51]. Reduced cognitive or attentional resources might hinder stable integration of articulatory patterns, and increased cognitive load could alter speech-motor control [6]. In the fiction and retell tasks, children with ADHD might allocate more cognitive resources to content generation than to articulation or prosody. The three tasks above fall into the category of *free speech*, highlighting the necessity of the inclusion of such tasks, as is already common in speech-and voice-based research [21].

Importantly, speech-based markers were evaluated in relation to an established clinical ADHD diagnosis and should therefore be interpreted as distinguishing clinically diagnosed from neurotypical children, rather than as stand-alone diagnostic markers. This study has several limitations. First, the modest sample size may have limited the detection of subtle effects, particularly in the linear regression analysis and correlations between voice features and symptom-scores. Second, recording equipment was updated during data collection; intensity-dependent features from the earlier setup were therefore excluded. Finally, as this was an observational study in children with ADHD, several aspects could not be standardised or monitored, like the time between visits, medication timing, or daily behaviour/feelings (like sleep or stress). Strengths of this study include a clinically well characterized ADHD cohort, most of whom were drug-naïve at baseline, and a well-matched control group with repeated assessments. Additionally, high-quality voice recordings were obtained in accordance with current guidelines and a standardised study protocol.

In summary, to our knowledge, this is the first clinical study to systematically examine task-dependent voice features and speech embeddings in children with ADHD, including their sensitivity to pharmacological treatment effects of MPH. The findings demonstrate that speech-based measures capture both disorder-related vocal characteristics and medication-associated change patterns in children with ADHD, with the picture description task being particularly informative.

## Supporting information

upporting_Information_Appendix_S1_S2_S3_S4_Tables_S1_S2

## Data Availability

The raw data underlying this study cannot be shared publicly due to the sensitive and potentially identifiable nature of voice recordings and associated clinical data. Where ethically and legally permissible, derived and non-identifiable summary features may be shared upon reasonable request and subject to data use agreements. We provide the full analysis code in a GitHub repository.

https://github.com/RaBa-fzj/adhd-voice-features

## Acknowledgements

The authors would like to thank the families and children/adolescents for their participation. They also thank Lena Siepe, Katharina Ebi and Franziska Laven for their valuable support to this study especially during data acquisition.

## Ethical Information

The study is in accordance with the Declaration of Helsinki, was approved by the *Ethics Committee of the Medical Faculty of RWTH Aachen University* (approval no. 032/20), and registered with the *German Clinical Trials Registry* (DRKS; registration no. DRKS00030766). Written informed consent was received from all parents and children, regardless of age.

## Contributions

R.B. and G.P. conceptualised the study. R.B., G.K., G.P. designed the SOPs for the voice assessments. R.B conducted the data acquisition with the help of research assistants. R.B., G.P., L.D.L, J.D. performed the statistical analysis. R.B. wrote the manuscript. All authors were involved in critically revising the manuscript, and all authors approved the final version.

## Data/Code availability

The raw data underlying this study cannot be shared publicly due to the sensitive and potentially identifiable nature of voice recordings and associated clinical data. Where ethically and legally permissible, derived and non-identifiable summary features may be shared upon reasonable request and subject to data use agreements. We provide the full analysis code in a GitHub repository (https://github.com/RaBa-fzj/adhd-voice-features)

