## Supplementary material for "Speech-Based Markers in Paediatric ADHD: A Longitudinal Case-Control Study of Voice Features and Medication Effects": upporting_Information_Appendix_S1_S2_S3_S4_Tables_S1_S2

### Supporting Information

#### Appendix S1

The Standard Operating Procedure (SOP) that was used to conduct the voice measurements/speech tasks was written and used in German. As a full translation is not possible, the corresponding author will be happy to provide the original document upon request.

#### Appendix S2

Within each participant, the same recording equipment was used for both visits.

Initial recording setup: Use of a cardioid microphone (*Elgato Wave:1*, 24-bit/48-kHz resolution, -25–15-dBFS sensitivity) connected to a laptop with a microphone-mouth distance between 30 and 40 cm. Recordings were made using either *Voice Memo*® (Apple, 2023) or *Audacity*® *Version 3.0* (Audacity, 2021).

Final recording setup: Use of an omnidirectional headset microphone (Sennheiser HSP 2 EW 3; frequency response: 20–20,000 Hz; sensitivity: 2 mV) positioned 2 cm from the corner of the mouth to ensure a constant recording distance for each child and measurement. The headset microphone was used with a preamplifier (*Focusrite Scarlett Solo 3rd Gen*; 20 Hz – 20 kHz ±0.1 dB frequency response, 24-bit/192-kHz resolution) and recordings were made using *Audacity*® *Version 3.0* (Audacity, 2021).

#### Appendix S3

For the frequency task the beginning of the first word and the end of the last word were detected, and the recording was cut according to these boundaries. For the picture description task, the retell and the fiction task the first word was detected, and the recording was cut to the desired length of 90 seconds (picture description) or 60 seconds (retell/fiction). For the phonation normal and loud tasks, the three sustained /a/ sound were detected, the middle 60% was cut out of each one (approximately 3 seconds) and these were concatenated to create a clip lasting approximately 9 second. Ambient noise recordings were checked for voice, after which they were cut 5 seconds in from the beginning.

### Appendix S4

The ambient noise level for measurements at a 30-cm distance should optimally be < 38 dB (non-weighted) (Patel et al., 2018). For the children who used a microphone the values in the present study ranged between 3.29 - 30.32 dB (non-weighted). With the headset microphone the maximum ambient noise level should be about 48–53 dB (non-weighted), which is considered acceptable for accurate soft voice measurements (Šrámková et al., 2015). As the children in the present study were not instructed to speak softly, we found all values acquired by the headset microphone to be acceptable, as they ranged between 41.92 - 53.82 dB (non-weighted). Values of Signal-to-Noise-Ratio (SNR) should ideally be > 10db for accurate measuring of the soundlevel in noise (Šrámková et al., 2015). SNR values measured in the present study, either by microphone or headset microphone, ranged between 20.46 - 38.95 dB.

### Appendix S5

A total of 54 children who met inclusion criteria were found eligible for the study. Seven of these children had missing data for various reasons: two children with ADHD dropped out after the first visit due to the termination of pharmacotherapy; two neurotypical controls were audibly hoarse due to colds at the second visit; and three children had technical issues at the first visit. Due to these issues, data from one case and one control had to be excluded entirely, and the loudness-sensitive features could not be analysed for one control that used a headset.

**Table S1** Information on voice measurements (Equipment, task, average length)

| Characteristic | Mean (SD) Length (s)* | ADHD Baseline (n) | ADHD Follow-Up (n) | Control Baseline (n) | Control Follow-Up (n) |
|---|---|---|---|---|---|
| **Equipment** | | | | | |
| Microphone | - | 9 | 9 | 1 | 1 |
| Headset | - | 17 | 16 | 25 | 24 |
| **Task Completion** | | | | | |
| Frequency | 9.8 (2.8) | 26 | 25 | 26 | 25 |
| Picture description | 89.5 (2.6) | 26 | 25 | 26 | 25 |
| Retell | 59.9 (0.6) | 24 | 23 | 26 | 25 |
| Fiction | 59.8 (0.7) | 25 | 25 | 26 | 25 |
| Phonation normal | 8.5 (1.5) | 20 | 19 | 26 | 25 |
| Phonation loud | 8.7 (1.4) | 17 | 19 | 26 | 25 |

***Note.*** Average length of the preprocessed audio recordings per task.

**Table S2** Information on pharmacotreatment of children with ADHD

| | Drug | Dose (mg) | Post-intake interval (hh:mm) |
|---|---|---|---|
| 1 | Equasym® Extended-release (ER) | 50 | 06:45 |
| 2 | Medikinet® ER | 20 | 05:30 |
| 3 | Equasym® ER | 60 | 06:25 |
| 4 | Equasym® ER | 60 | 06:15 |
| 5 | Equasym® ER | 40 | 02:45 |
| 6 | Equasym® ER | 40 | 02:35 |
| 7 | Medikinet® ER | 10 | 02:35 |
| | Kinecteen® Immediate-release (IR) | 54 | |
| 8 | Kinecteen® ER | 36 | 04:05 |
| 9 | Medikinet® ER | 20 | 04:30 |
| 10 | Medikinet® ER | 10 | 02:30 |
| 11 | Equasym® ER | 30 | 02:50 |
| 12 | Concerta® ER | 36 | 02:15 |
| 13 | Equasym® ER | 30 | 03:50 |
| 14 | Equasym® ER | 20 | 04:00 |
| 15 | Medikinet® ER | 40 | 03:30 |
| 16 | Kinecteen® ER | 40 | 04:00 |
| 17 | Equasym® ER | 30 | 03:50 |
| 18 | Equasym® ER | 20 | 03:20 |
| 19 | Ritalin LA® IR | 40 | 03:50 |
| 20 | Medikinet® ER | 30 | 04:20 |
| 21 | Ritalin LA® ER | 30 | 03:30 |
| 22 | Equasym® ER | 30 | 03:30 |
| 23 | Equasym® ER | 30 | 03:30 |
| 24 | Medikinet® ER | 15 | 03:00 |
| 25 | Ritalin LA® ER | 30 | 03:20 |
| | | Average post-intake interval (SD) | 03:51 (01:13) |

***Note.*** One participant was given Medikinet® and Kinecteen®at the same time. Patients 1-22 were initially drug-naïve; 23 - 25 took a break from medication lasting ≥ 4 weeks before the follow-up. Post-intake interval is the time between taking the drug and the start of the speech assessment.
